# Ethnicity and clinical outcomes in COVID-19: A Systematic Review and Meta-analysis

**DOI:** 10.1101/2020.09.05.20188821

**Authors:** Shirley Sze, Daniel Pan, Laura J Gray, Clareece R Nevill, Christopher A Martin, Joshua Nazareth, Jatinder S Minhas, Pip Divall, Kamlesh Khunti, Keith R Abrams, Laura B Nellums, Manish Pareek

**Affiliations:** Department of Cardiovascular Sciences, University of Leicester, United Kingdom; Department of Respiratory Sciences, University of Leicester, United Kingdom; Department of Infection and HIV Medicine, University Hospitals Leicester NHS Trust, United Kingdom; Department of Health Sciences, University of Leicester, United Kingdom; University Hospitals of Leicester, Education Centre Library, Glenfield Hospital and Leicester Royal Infirmary, United Kingdom; Diabetes Research Centre, University of Leicester, United Kingdom; Division of Epidemiology and Public Health, School of Medicine, University of Nottingham, United Kingdom

## Abstract

**Importance:** The association of ethnicity with outcomes in patients with COVID-19 is unclear.

**Objective:** To determine whether the risk of SARS-CoV-2 infection, COVID-19 intensive care unit (ICU) admission and mortality are associated with ethnicity.

**Data Sources:** We searched all English language articles published 1^st^ December 2019 - 30^th^ June 2020 within MEDLINE, EMBASE, PROSPERO and the Cochrane library using indexing terms for COVID-19 and ethnicity, as well as manuscripts awaiting peer review on MedRxiv during the same period.

**Study Selection:** Included studies reported original clinical data, disaggregated by ethnicity, on patients with confirmed or suspected COVID-19. We excluded correspondence, area level, modelling and basic science articles. Two independent reviewers screened articles for inclusion. Of 926 identified articles, 35 were included in the meta-analyses.

**Data Extraction and Synthesis:** The review was conducted according to PRISMA guidelines. Reviewers independently extracted data using a piloted form on: (1) rates of infection, ICU admission and mortality by ethnicity; and (2) unadjusted and adjusted data comparing ethnic minority and White groups. Data were pooled using random effects models.

**Main Outcomes and Measures:** Outcomes were: (1) infection with SARS-CoV-2 confirmed on molecular testing; (2) ICU admission; and (3) mortality in COVID-19 confirmed and suspected cases.

**Results:** 13,535,562 patients from 35 studies were included in the meta-analyses. Black, Asian and Hispanic individuals had a greater risk of infection compared to White individuals (Black: pooled adjusted RR: 2.06, 95% CI: 1.59-2.67; Asian: 1.35, 95%CI: 1.15-1.59; Hispanic: 1.77, 95% CI: 1.39-2.25). Black individuals were significantly more likely to be admitted to ICU than White individuals (pooled adjusted RR: 1.61, 95% CI: 1.02-2.55). Risk of mortality was similar across ethnicities among hospitalised patients, but increased among Asian and Mixed ethnic groups in the general population.

**Conclusions:** Black, Asian and Hispanic ethnic groups are at increased risk of SARS-CoV-2 infection. Black individuals may be more likely to require ICU admission for COVID-19. There may also be disparities in risk of death from COVID-19 at a population level. Our findings are of critical public health importance and should inform policy on minimising SARS-CoV-2 exposure in ethnic minority groups.

**KEY POINTS:** *Question:* Is ethnicity associated with vulnerability to, and outcomes from, coronavirus disease 2019 (COVID-19)?

*Findings:* In this systematic review and meta-analysis, rates of infection and outcomes from COVID-19 were compared between ethnic groups. Individuals from Black, Asian and Hispanic ethnicity were significantly more vulnerable to SARS-CoV-2 infection than those of White ethnicity. Black individuals were more likely to need intensive care unit (ICU) admission for COVID-19 than White individuals. Risk of mortality was similar across ethnicities among hospitalised patients, but increased among Asian and Mixed ethnic groups in the general population.

*Meaning:* There is strong evidence for an increased risk of SARS-CoV-2 infection amongst ethnic minorities, and targeted public health policies are required to reduce this risk.

## Introduction

Over 25 million people have been infected with severe acute respiratory virus-2 (SARS-CoV-2) since December 2019.^1^ Race and ethnicity has come under scrutiny as an important risk factor for infection, severe disease and death, with evidence that ethnic minorities may be at increased risk of COVID-19 morbidity and mortality.^2,3^

Understanding the relationship between ethnicity and COVID-19 is an urgent research priority, in order to reduce the disproportionate burden of disease in these populations.^4,5^ Recently there has been an increase in the volume of literature, both published and in preprint servers, on the association of ethnicity with vulnerability to COVID-19 infection and clinical outcomes.^6^ A comprehensive synthesis of existing evidence examining the relationship between ethnicity and COVID-19 is urgently needed in order to inform clinical care and public health policy. We therefore conducted a systematic review and meta-analysis of both published and preprint research to study the association of ethnicity with COVID-19 infection and outcomes. Specifically, we aimed to identify ethnic differences in the risk of becoming infected with SARS-CoV-2 as well as subsequent intensive care admission and death.

## Methods

We conducted the research according to the Preferred Reporting Items for Systematic Reviews and Meta-Analyses (PRISMA) guidelines, and registered our review on PROSPERO (CRD42020180654).^7^

### Data sources and searches

An academic librarian (PD) developed the search strategies, and carried out a search of the databases MEDLINE, EMBASE, PROSPERO and the Cochrane Library, as shown in the Supplementary materials 1. We searched for any articles published in English from 1^st^ December 2019 to 30^th^ June 2020. We also reviewed any publications awaiting peer review on MedRxiv during the same period.

### Eligibility criteria

We included studies with original clinical data on COVID-19 infection, intensive care admission, and mortality disaggregated by ethnicity. We excluded correspondence pieces, area level studies (reporting aggregated data rather than individual risk), and predictive modelling studies, or those that only included basic science or animal data or did not report population data (e.g. studies of infection breakouts). Retracted papers were also excluded. If studies assessed race and ethnicity separately, data were only extracted for mutually exclusive groups. Where multiple preprints of the same database have been published (for example, the Biobank database), the most recent version up to 30^th^ June 2020 was used, with published peer-reviewed studies favoured over those in the preprint database. Studies which assessed two different cohorts in the same paper were included as two separate datasets in the analysis. We used rigorous methods to exclude duplicate publications by identifying centres and excluding studies using the same database.

### Study selection

Using a standardised piloted form, DP and SS independently screened titles, abstracts and full-text articles reporting potentially eligible studies. Disagreements were resolved by discussion or consultation with an adjudicator (MP) when necessary.

### Data extraction

One reviewer (DP, SS, CAM, JN, JSM, LBN) independently extracted data from each article. Data extraction was duplicated for all papers by an independent researcher (CN). We stratified patients into the following ethnic groups based on the categorisations used in the included papers: White; Asian (including South Asian, Asian/Pacific-Islander and Chinese); Black; Hispanic; Native American; Mixed and Other. When the proportion of patients of each ethnicity was not presented in the text, we calculated the proportion from data in the manuscript.

The outcomes studied were:

- Infection with SARS-CoV-2
- Intensive Care Unit (ICU) admission
- Death

Patients with COVID-19 were defined as those who tested positive for SARS-CoV-2 by nasopharyngeal swab or had clinical symptoms and signs of COVID-19.

### Data synthesis and analysis

We first synthesised prevalence of each outcome and unadjusted data by ethnicity. We excluded studies which did not record any events. Raw counts were used for the unadjusted data to calculate odds ratios (OR) and 95% confidence intervals (CI).

We also synthesised data adjusted for key confounders. For risk of infection and ICU admission, we extracted adjusted risk ratios (RR). Adjusted OR were converted to adjusted RR using the conversion method as recommended by the Cochrane Handbook.^8^ For mortality, we extracted adjusted hazard ratios (HR) (95% CI) where possible, and assumed adjusted RR to approximate an adjusted HR.

Some studies presented multiple models with different sets of confounders. We included the model which most closely matched our a priori chosen confounders of age, sex, deprivation, obesity, and comorbidities. For both the adjusted and unadjusted comparisons, data were extracted for analyses which used White ethnicity as the reference group.

For ICU admission and mortality, we included studies which reported suspected or confirmed COVID-19 patients in their analyses. For mortality, we conducted a further analysis to include studies which looked at the risk of death from COVID-19 in the general population (i.e., those with and without COVID-19). Sensitivity analyses were also conducted excluding studies which did not include data for those still hospitalised at the end of the study, and studies which were of mixed populations (hospitalised and non-hospitalised patients).

For all outcomes and data types, we synthesised data (prevalence, unadjusted OR and adjusted RR/HR) using the DerSimonian and Laird random effects model.^9^ I^2^ was used to assess heterogeneity. All meta-analyses were conducted using STATA version 16.1 (StataCorp, United States). P values < 0.05 were considered to be statistically significant.

### Quality of evidence

Joanna Briggs Institute Critical Appraisal Tools were used to assess the quality of evidence for all studies relevant to each study design.^10^ Publication bias was assessed visually using funnel plots and formally with Egger’s test for analyses including at least 10 studies.^11^

## Results

### Study selection

282 articles were identified in the published literature between 1^st^ December 2019 and 30^th^ June 2020 as shown in Figure 1. An additional 644 articles were identified from MedRxiv within the same period, giving a total of 926 articles. 126 articles were excluded because they did not report on individuals with COVID-19; 57 were excluded due to no mention of ethnicity. Two papers were excluded due to retraction during the study period. Of the remaining 741 articles that were assessed for eligibility in full-text screening, 638 did not meet inclusion criteria (reasons for exclusion, Figure 1). A further 68 were excluded from the meta-analysis, including 62 studies which provided no data on our predefined outcomes, and six studies which used the same database as other included studies.

**Figure 1:**
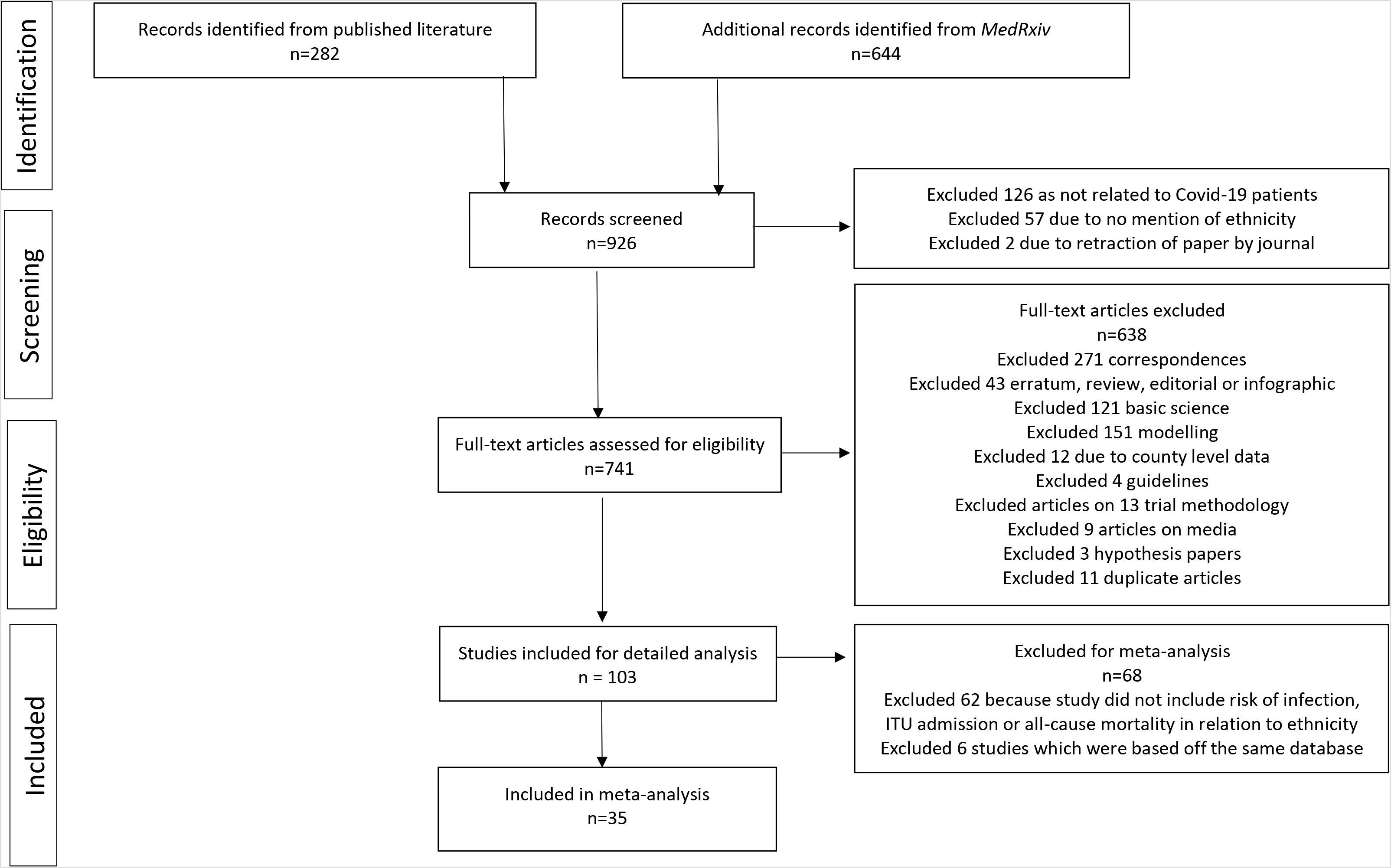
PRISMA flowchart of study

### Study characteristics

A total of 13,535,562 patients from 35 papers^12–45^ were included in the meta-analyses; 11,383,563 (84%) were White; 1,045,656 (8%) were Asian; 383,970 (2.8%) were Black, 32,066 (0.2%) were Hispanic, two were Native American, 349,038 (2.5%) were Mixed, and 341,267 (2.5%) were of Other ethnic group.

Detailed descriptions of the included studies are shown in Table 1. Most studies (n = 28, 80%) were from the USA; the remaining six (17%) were from the UK. One study (3%) described two separate cohorts from the USA and the UK.^19^ One study (3%) was a case series; two (6%) were cross-sectional and the remaining (91%) were cohort studies. 20 (57%) reported on patients in hospital; three (9%) reported on patients in the community; 12 (34%) reported on both. As of 31^st^ August 2020, half (n = 18, 51%) of papers included were published.

**Table 1:**
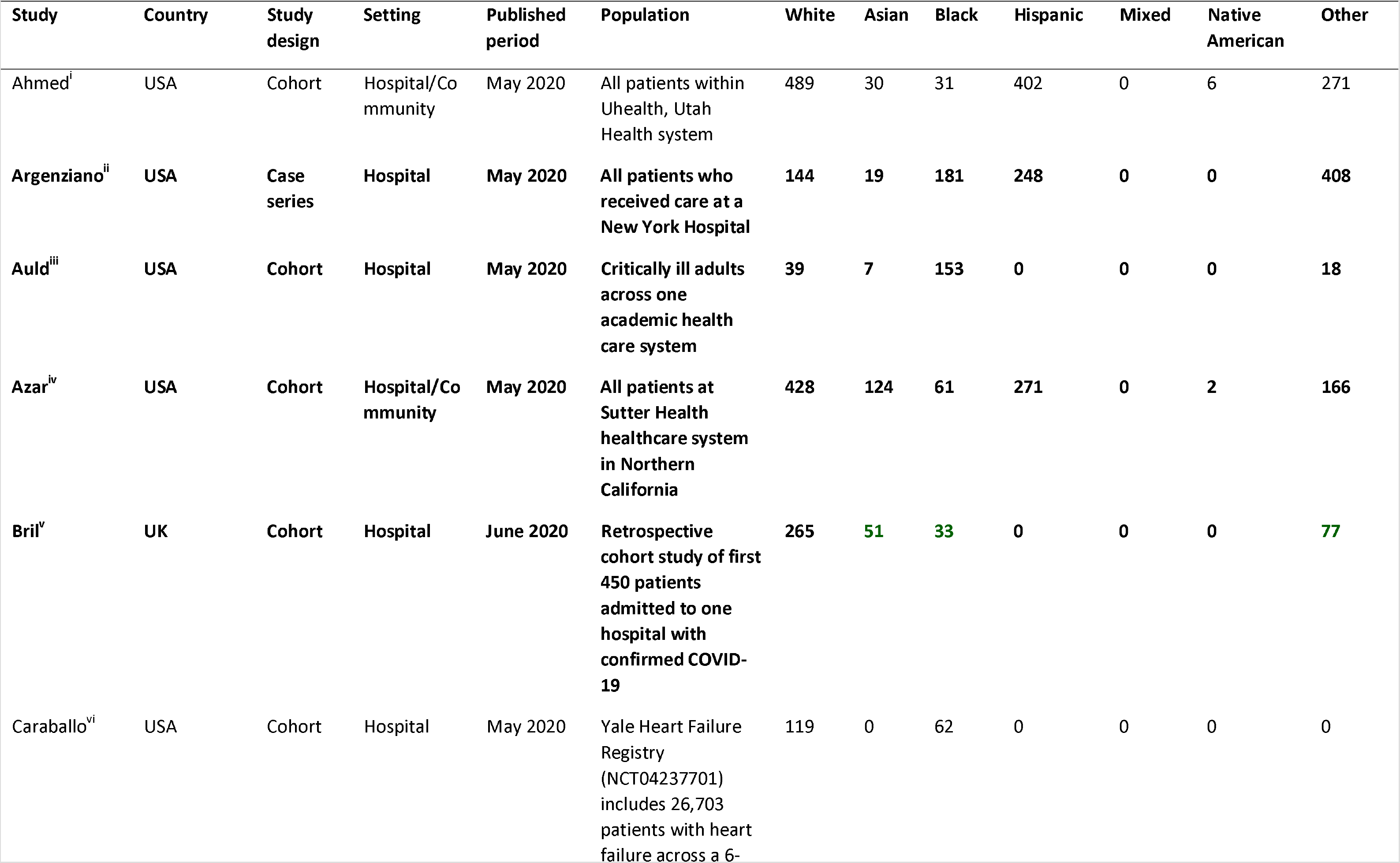
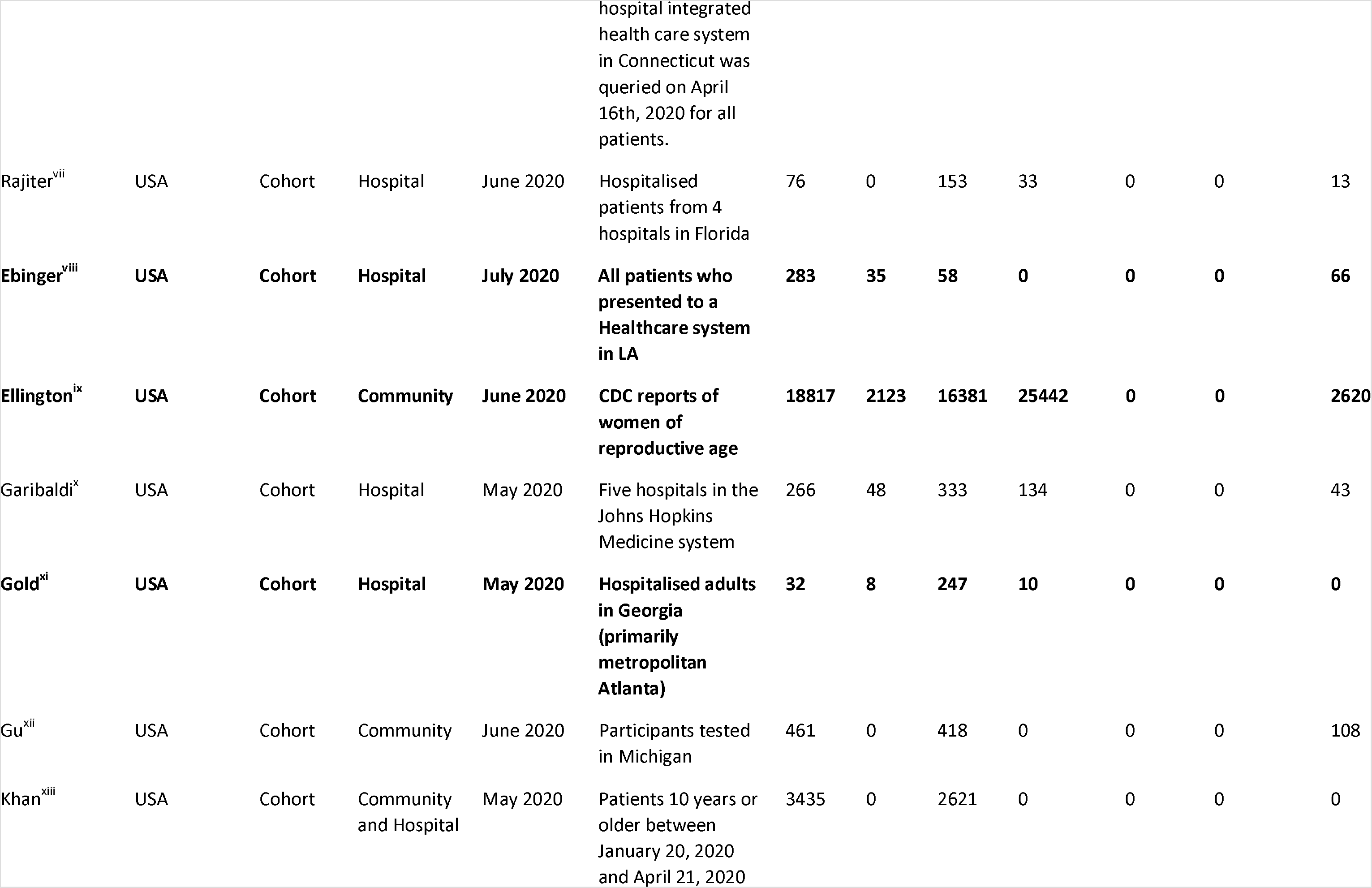
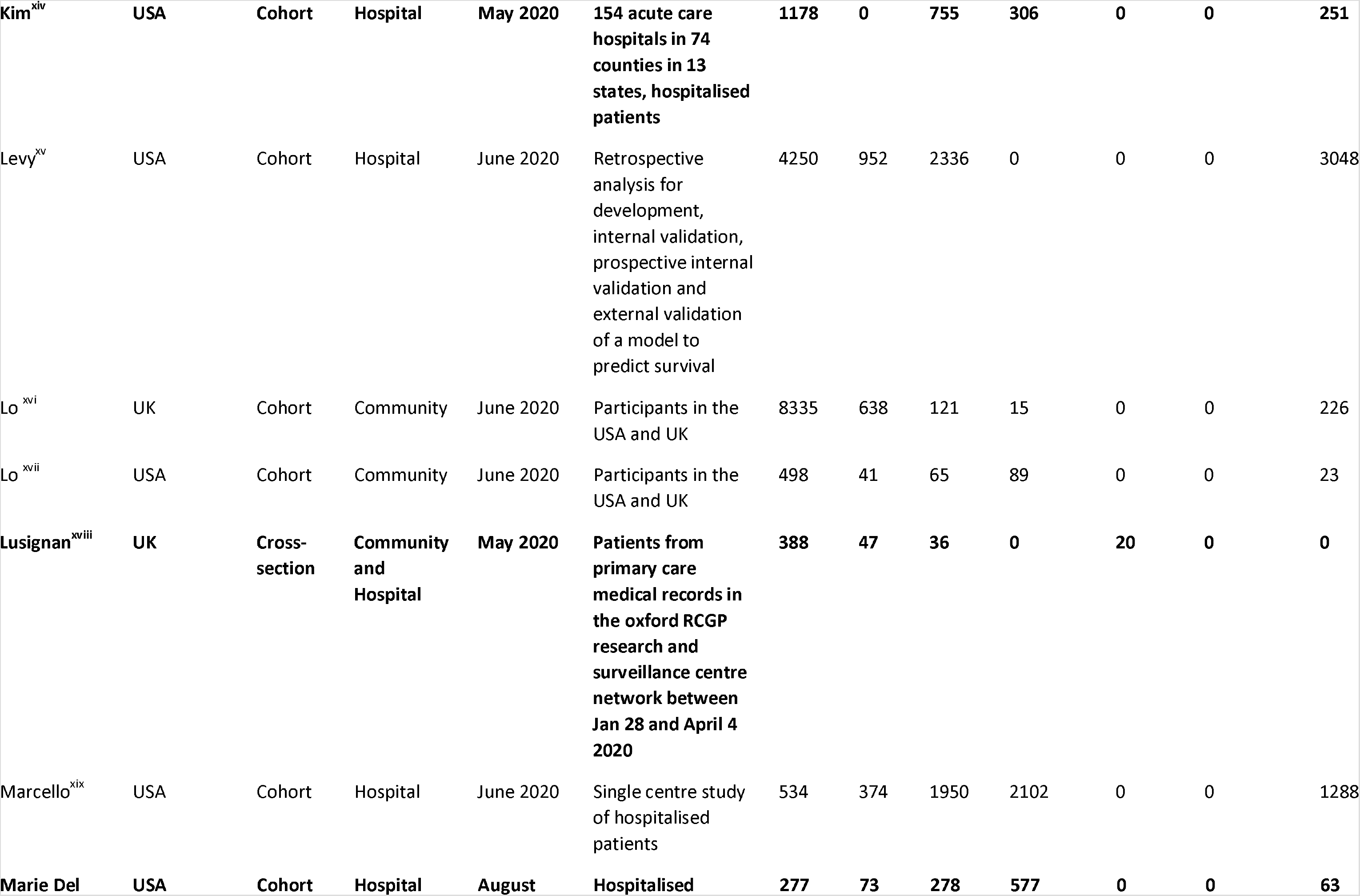
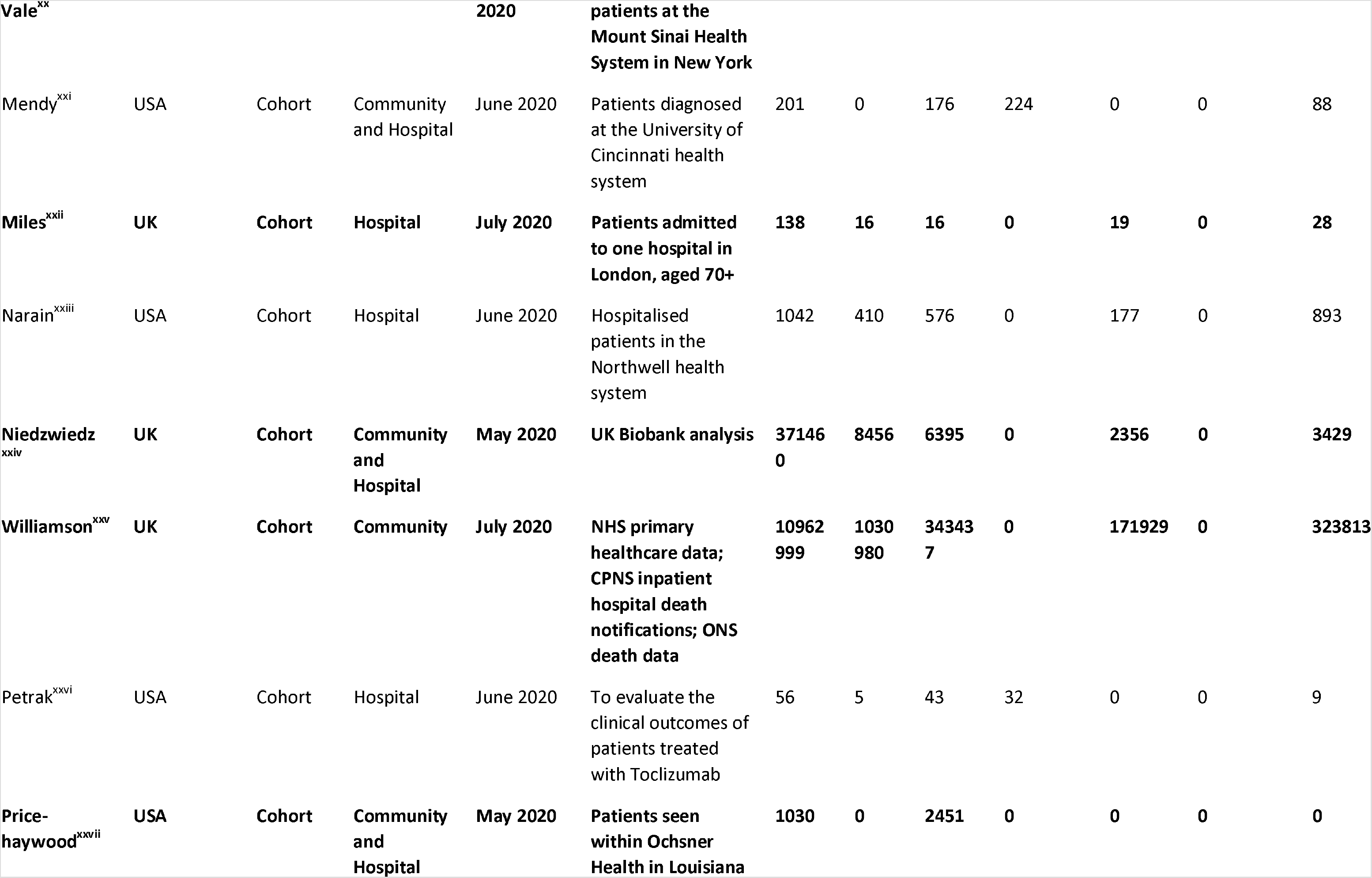
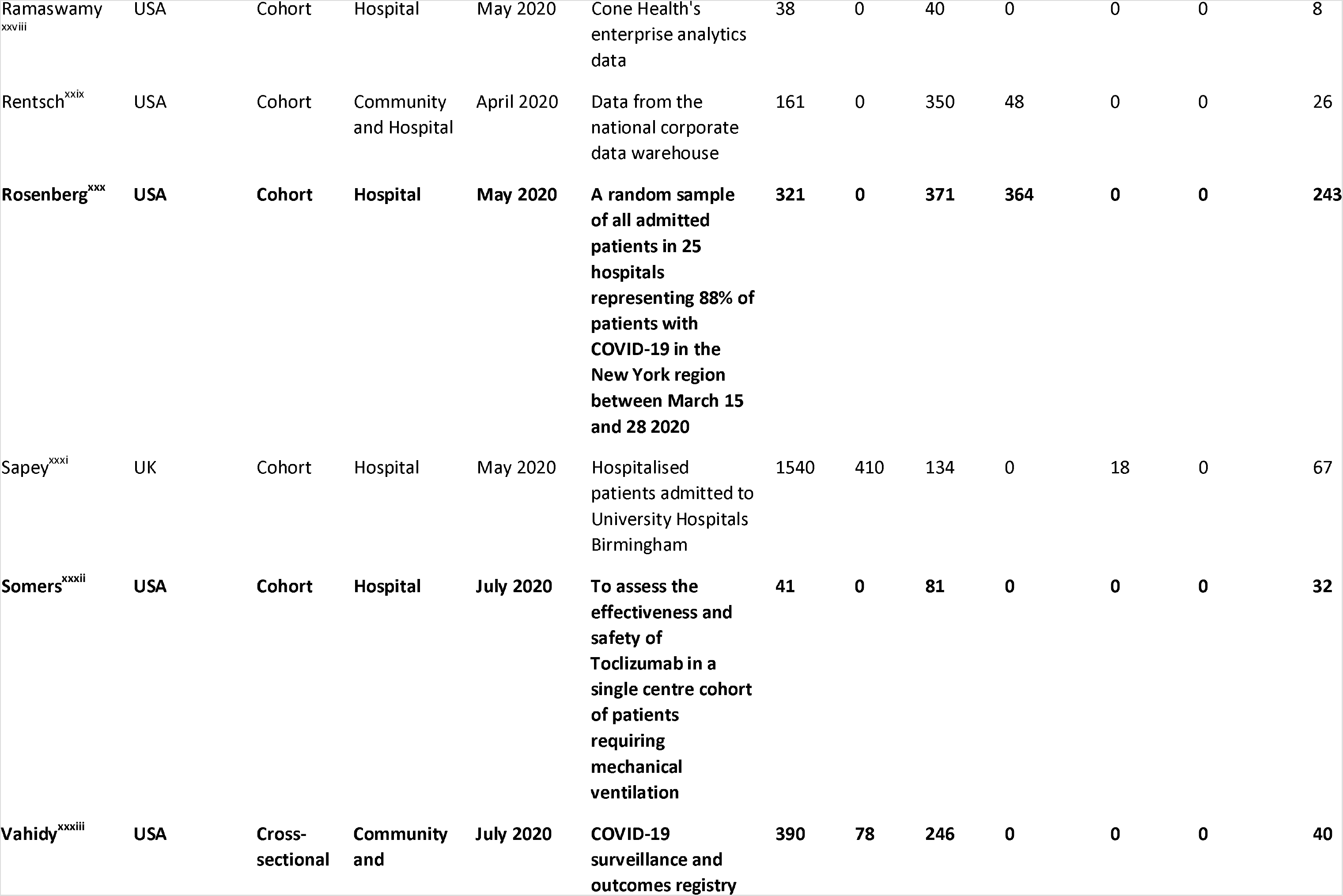
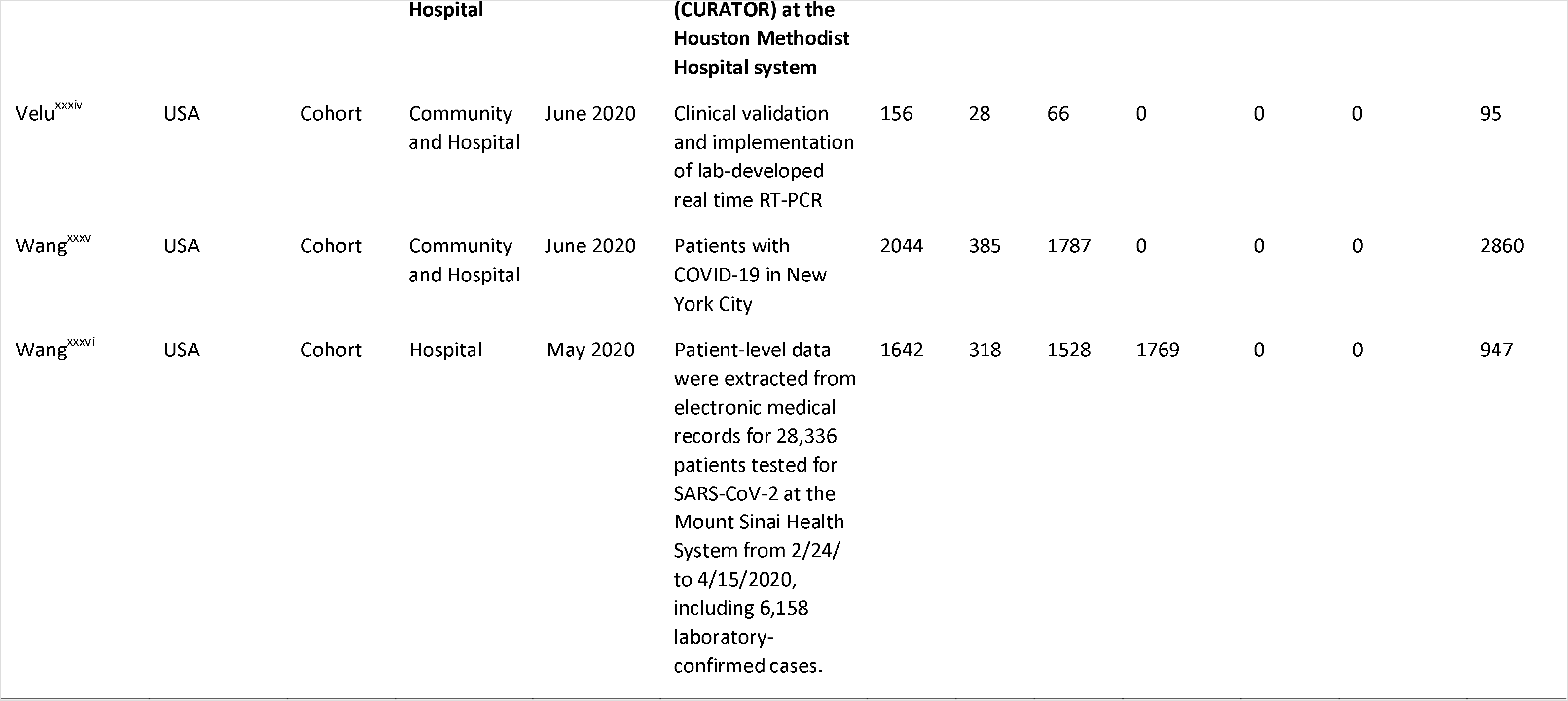
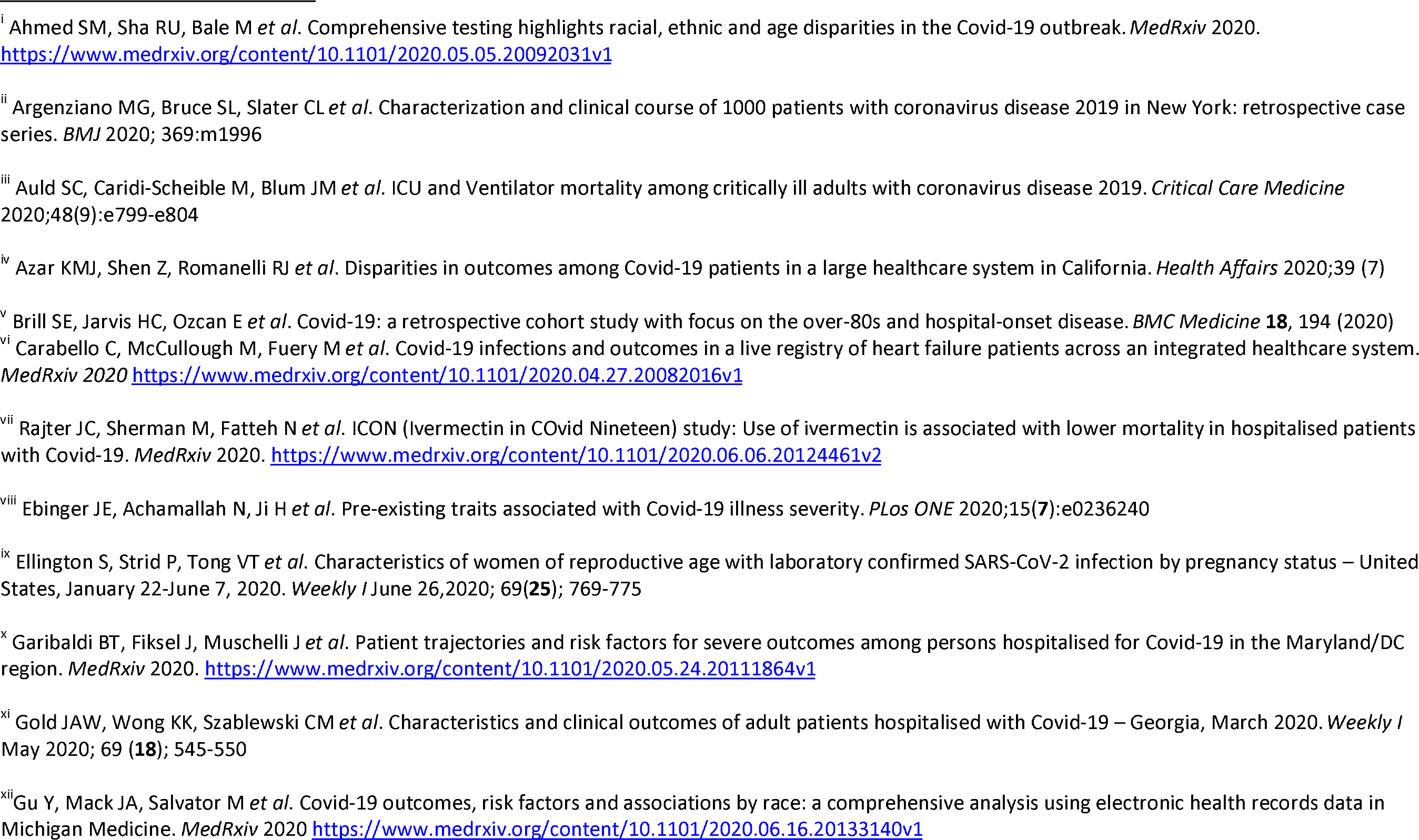
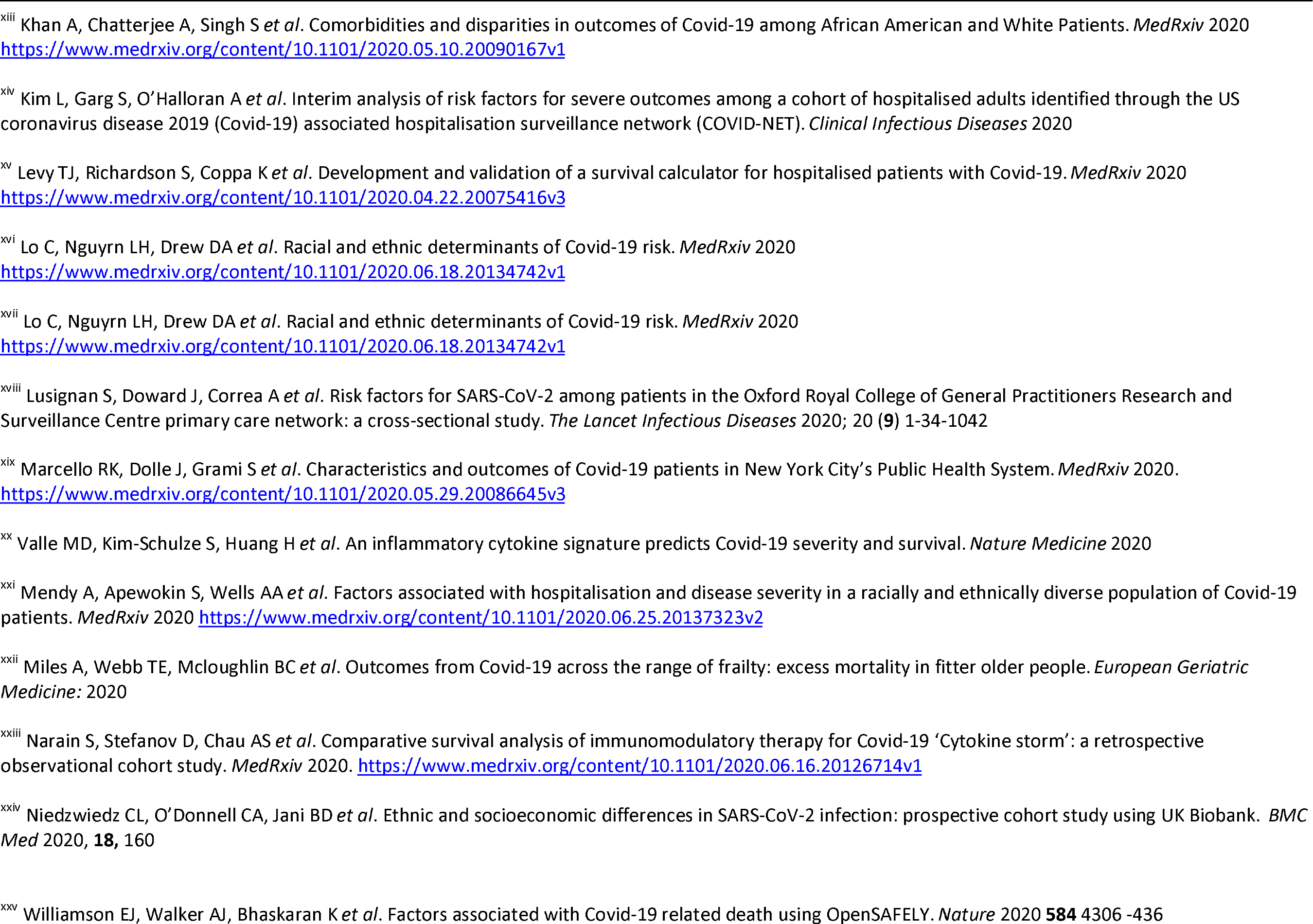
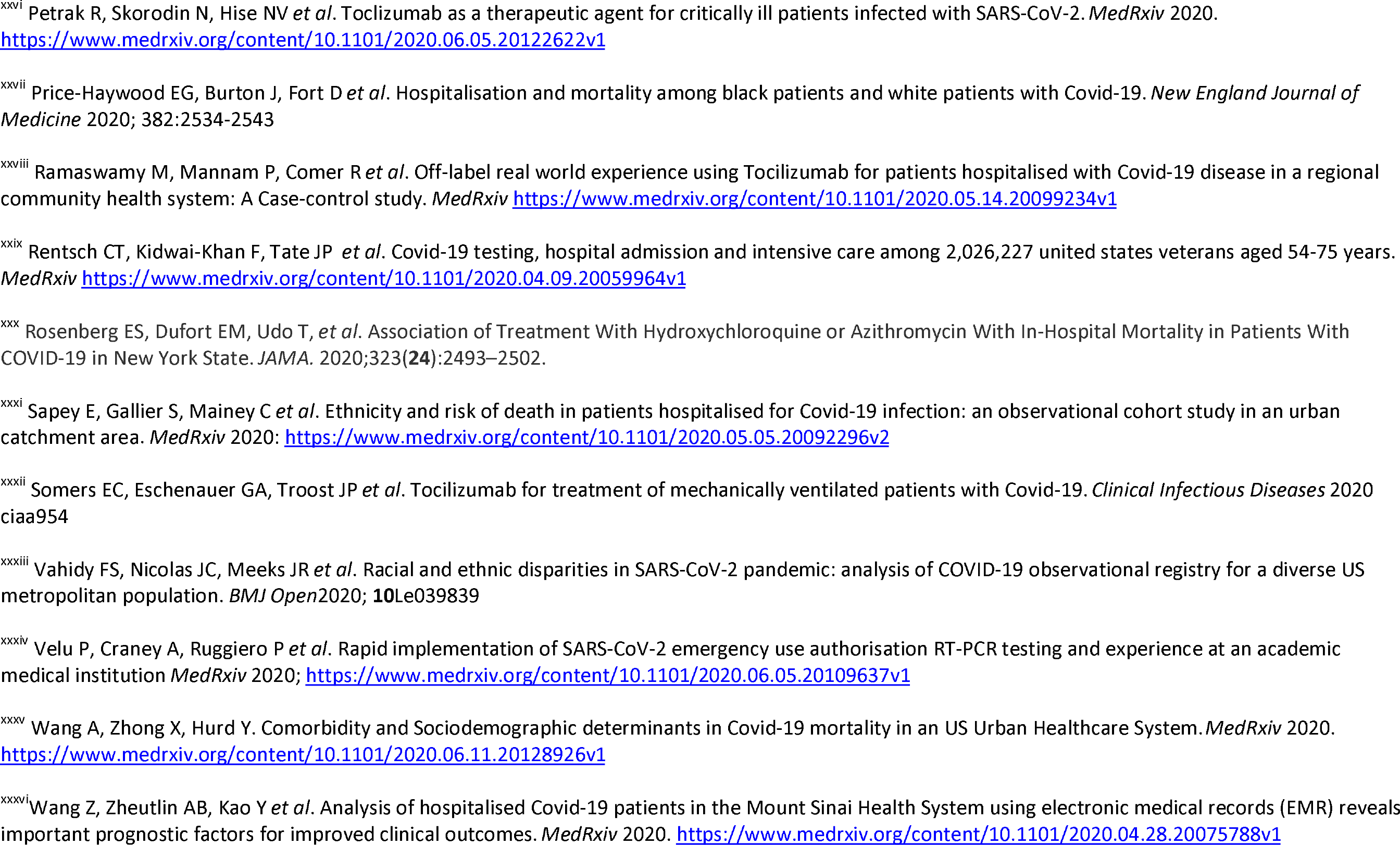
Characteristics of the 35 included studies. Rows in bold are articles which have been published in peer reviewed journals.

**Table 2:** Data syntheses for risk of infectivity of SARS-CoV-2 by differing ethnic groups.

|  | Studies | Pooled prevalence<br>(95% CI) | I<sup>2</sup> | Studies | Pooled OR<br>(95% CI) | I<sup>2</sup> | Studies | Pooled RR<br>(95% CI) | I<sup>2</sup> |
| --- | --- | --- | --- | --- | --- | --- | --- | --- | --- |
| White | 13 | 0.13 (0.12, 0.13) | 99.9 |  | Reference |  |  | Reference |  |
| Asian | 10 | 0.18 (0.15, 0.20) | 99.6 | 10 | **1.55 (1.13, 2.13)** | 96.3 | 4 | **1.35 (1.15, 1.59)** | 0 |
| Black | 13 | 0.29 (0.22, 0.36) | 99.9 | 12 | **2.50 (1.88, 3.32)** | 96.6 | 6 | **2.06 (1.59, 2.67)** | 86.7 |
| Hispanic | 7 | 0.21 (0.09, 0.33) | 99.97 | 7 | **2.44 (1.90, 3.15)** | 94.8 | 3 | **1.77 (1.39, 2.25)** | 0 |
| Mixed | 1 | 0.33 (0.16, 0.56) | - | 1 | 0.96 (0.36, 2.58) | - | 0 | - | - |
| Native American | 1 | 0.02 (0.01, 0.07) | - | 1 | 0.31 (0.08, 1.26) | - | 0 | - | - |

The overall quality of published articles was higher than those in preprint; their average rating was 82% compared to 75% for preprint articles as shown in Table 1.

### Risk of SARS-CoV-2 infection

Thirteen studies reported data on the risk of SARS-CoV-2 infection in those tested stratified by ethnicity, as shown in Table 2 and Supplementary materials 2. Pooled prevalence was highest in Mixed and Black groups. After adjusting for confounders, individuals of Black ethnicity had double the risk of SARS-CoV-2 compared to those of White ethnicity (pooled adjusted RR: 2.06, 95% CI: 1.59-2.67). There was also an increased risk of SARS-CoV-2 infection among those of Asian (pooled adjusted RR: 1.35, 95% CI: 1.15-1.59) and Hispanic ethnicity (pooled adjusted RR: 1.77, 95% CI: 1.39-2.25) compared to White ethnicity. Data for Mixed and Native American ethnicity were limited by small numbers of patients. Across all pooled analyses, there were high levels of heterogeneity.

**Table 3:** Data syntheses for risk of ICU admission amongst different ethnic groups.

|  | Studies | Pooled prevalence<br>(95% CI) | I<sup>2</sup> | Studies | Pooled unadjusted<br>OR<br>(95% CI) | I<sup>2</sup> | Studies | Pooled adjusted<br>RR<br>(95% CI) | I<sup>2</sup> |
| --- | --- | --- | --- | --- | --- | --- | --- | --- | --- |
| **(1) Studies including both hospitalised and inpatient/outpatient populations** |  |  |  |  |  |  |  |  |  |
| White | 10 | 0.15 (0.10, 0.20) | 98.1 |  | Reference |  |  | Reference |  |
| Asian | 5 | 0.16 (0.09, 0.24) | 86.5 | 5 | 1.35 (0.69, 2.64) | 85.0 | 0 | - | - |
| Black | 11 | 0.24 (0.19, 0.28) | 94.9 | 10 | **1.73 (1.17, 2.56)** | 92.7 | 4 | **1.61 (1.02, 2.55)** | 92.1 |
| Hispanic | 6 | 0.17 (0.11, 0.23) | 93.0 | 6 | 0.88 (0.71, 1.10) | 41.3 | 2 | 1.62 (0.53, 4.97) | 88.5 |
| Native American | 0 | - | - | 1 | 1.65 (0.08, 34.79) | - | 0 | - | - |
| **(2) Studies including hospitalised populations only or reporting a subgroup analysis for hospitalised patients only** |  |  |  |  |  |  |  |  |  |
| White | 10 | 0.28 (0.21, 0.36) | 97.9 |  | Reference |  |  | Reference |  |
| Asian | 5 | 0.27 (0.17, 0.36) | 80.4 | 5 | 1.68 (0.95, 2.99) | 76.2 | 0 | - | - |
| Black | 11 | 0.35 (0.25, 0.44) | 97.8 | 10 | **1.29 (1.06, 1.58)** | 61.9 | 1 | 1.01 (0.89, 1.15) | - |
| Hispanic | 6 | 0.31 (0.19, 0.42) | 96.1 | 6 | 0.97 (0.71, 1.33) | 66.5 | 1 | 0.96 (0.76, 1.21) | - |

### ICU admission

Eleven studies reported data on the risk of ICU admission by ethnicity among COVID-19 patients, as shown in Table 3 and Supplementary materials 3. Pooled prevalence was highest in the Black group. Of these studies, four provided comparative estimates adjusted for confounders. From the adjusted pooled estimates, individuals of Black and Hispanic ethnicity were more likely to be admitted to ICU compared to those of White ethnicity. Although the effect estimates were similar (Black: pooled adjusted RR: 1.61, 95% CI: 1.02-2.55; Hispanic: pooled adjusted RR: 1.62, 95% CI 0.53-4.97), the association was only significant for those of Black ethnicity. Only unadjusted data were available for comparing the risk of ICU admission in those of Asian ethnicity to White ethnicity. This showed a trend towards increased risk of ICU admission (OR: 1.68, 95% CI: 0.95-2.99). One study presented data for Native American ethnicity (OR: 1.54, 95% CI: 0.08-34.79) and no studies for Mixed ethnicity.^35^ Across all pooled analyses, there were high levels of heterogeneity.

Sensitivity analysis limited to hospitalised patients showed a higher prevalence of ICU admission for all ethnic groups compared to studies reporting hospitalised and community populations combined. Only one study presented adjusted RR for Black and Hispanic ethnic groups compared to White ethnicity for hospitalised patients, which showed no increased risk for ICU admission.^17^

### Mortality

Twenty three studies provided data on the risk of mortality in patients with COVID-19, by ethnic groups, as shown in Table 4 and Supplementary materials 4. Pooled prevalence was highest in Asian and Mixed groups. Eleven studies provided adjusted comparative data; four provided adjusted data, but did not provide raw counts or unadjusted comparative data. An additional two studies provided data on the risk of mortality in COVID-19 patients in the general population, and were included in our sensitivity analysis.

**Table 4:**
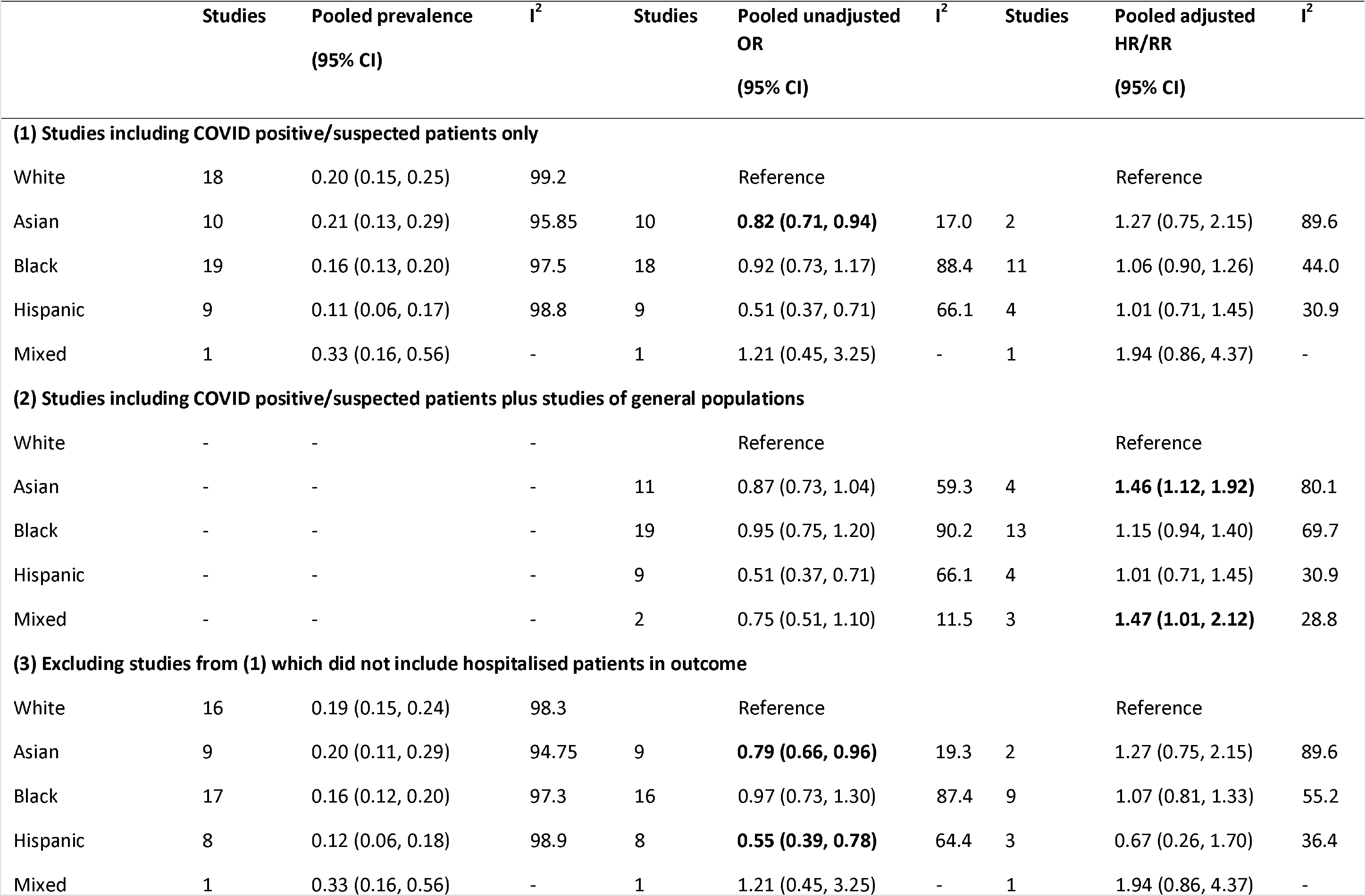
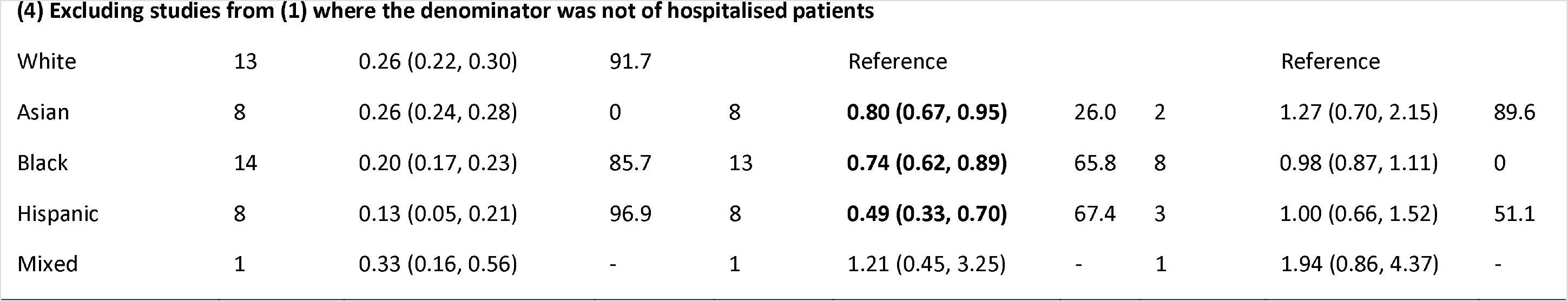
Data syntheses for risk of mortality by different ethnic groups.

|  | Studies | Pooled prevalence<br>(95% CI) | I<sup>2</sup> | Studies | Pooled unadjusted<br>OR<br>(95% CI) | I<sup>2</sup> | Studies | Pooled adjusted<br>HR/RR<br>(95% CI) | I<sup>2</sup> |
| --- | --- | --- | --- | --- | --- | --- | --- | --- | --- |
| **(1) Studies including COVID positive/suspected patients only** |  |  |  |  |  |  |  |  |  |
| White | 18 | 0.20 (0.15, 0.25) | 99.2 |  | Reference |  |  | Reference |  |
| Asian | 10 | 0.21 (0.13, 0.29) | 95.85 | 10 | **0.82 (0.71, 0.94)** | 17.0 | 2 | 1.27 (0.75, 2.15) | 89.6 |
| Black | 19 | 0.16 (0.13, 0.20) | 97.5 | 18 | 0.92 (0.73, 1.17) | 88.4 | 11 | 1.06 (0.90, 1.26) | 44.0 |
| Hispanic | 9 | 0.11 (0.06, 0.17) | 98.8 | 9 | 0.51 (0.37, 0.71) | 66.1 | 4 | 1.01 (0.71, 1.45) | 30.9 |
| Mixed | 1 | 0.33 (0.16, 0.56) | - | 1 | 1.21 (0.45, 3.25) | - | 1 | 1.94 (0.86, 4.37) | - |
| **(2) Studies including COVID positive/suspected patients plus studies of general populations** |  |  |  |  |  |  |  |  |  |
| White | - | - | - |  | Reference |  |  | Reference |  |
| Asian | - | - | - | 11 | 0.87 (0.73, 1.04) | 59.3 | 4 | **1.46 (1.12, 1.92)** | 80.1 |
| Black | - | - | - | 19 | 0.95 (0.75, 1.20) | 90.2 | 13 | 1.15 (0.94, 1.40) | 69.7 |
| Hispanic | - | - | - | 9 | 0.51 (0.37, 0.71) | 66.1 | 4 | 1.01 (0.71, 1.45) | 30.9 |
| Mixed | - | - | - | 2 | 0.75 (0.51, 1.10) | 11.5 | 3 | **1.47 (1.01, 2.12)** | 28.8 |
| **(3) Excluding studies from (1) which did not include hospitalised patients in outcome** |  |  |  |  |  |  |  |  |  |
| White | 16 | 0.19 (0.15, 0.24) | 98.3 |  | Reference |  |  | Reference |  |
| Asian | 9 | 0.20 (0.11, 0.29) | 94.75 | 9 | **0.79 (0.66, 0.96)** | 19.3 | 2 | 1.27 (0.75, 2.15) | 89.6 |
| Black | 17 | 0.16 (0.12, 0.20) | 97.3 | 16 | 0.97 (0.73, 1.30) | 87.4 | 9 | 1.07 (0.81, 1.33) | 55.2 |
| Hispanic | 8 | 0.12 (0.06, 0.18) | 98.9 | 8 | **0.55 (0.39, 0.78)** | 64.4 | 3 | 0.67 (0.26, 1.70) | 36.4 |
| Mixed | 1 | 0.33 (0.16, 0.56) | - | 1 | 1.21 (0.45, 3.25) | - | 1 | 1.94 (0.86, 4.37) | - |
| **(4) Excluding studies from (1) where the denominator was not of hospitalised patients** |  |  |  |  |  |  |  |  |  |
| White | 13 | 0.26 (0.22, 0.30) | 91.7 |  | Reference |  |  | Reference |  |
| Asian | 8 | 0.26 (0.24, 0.28) | 0 | 8 | **0.80 (0.67, 0.95)** | 26.0 | 2 | 1.27 (0.70, 2.15) | 89.6 |
| Black | 14 | 0.20 (0.17, 0.23) | 85.7 | 13 | **0.74 (0.62, 0.89)** | 65.8 | 8 | 0.98 (0.87, 1.11) | 0 |
| Hispanic | 8 | 0.13 (0.05, 0.21) | 96.9 | 8 | **0.49 (0.33, 0.70)** | 67.4 | 3 | 1.00 (0.66, 1.52) | 51.1 |
| Mixed | 1 | 0.33 (0.16, 0.56) | - | 1 | 1.21 (0.45, 3.25) | - | 1 | 1.94 (0.86, 4.37) | - |

From the adjusted pooled estimates of studies which looked at hospitalised patients with COVID-19, patients from ethnic minority backgrounds have a similar risk of death compared to those of White ethnicity. Sensitivity analysis which investigated the risk of death from COVID-19 in the general population showed that patients of Asian (pooled adjusted HR 1.46, 95% CI: 1.12-1.92) and Mixed (pooled adjusted HR: 1.47, 95% CI: 1.01-2.12) ethnicities have an increased risk of death compared to those of White ethnicity. Further sensitivity analyses excluding (1) studies not reporting mortality data on patients who were still hospitalised at the end of the study and (2) studies which were of mixed ethic populations showed that patients from ethnic minority groups had a similar risk of death compared to those of White ethnicity.

### Risk of bias across studies

For prevalence estimates of infection, there was clear asymmetry in funnel plots for Asian, Black and White ethnic groups (Egger’s test *p* = 0.003, *p* = 0.007 and *p* = 0.001 respectively, Supplementary materials 5). Studies with very low estimates of infection had very high precision, whereas studies with higher infection estimates had lower precision.

For prevalence estimates of ICU admission, there was some asymmetry in the funnel plot for White ethnic group (Egger’s test *p* = 0.04). For prevalence estimates of mortality, there was some asymmetry in the funnel plot for the Black ethnic group (Egger’s test p = 0.006). The asymmetry appears to be driven by small-study effects; the studies with low precision all had higher estimates of prevalence.

## DISCUSSION

We present the first meta-analysis investigating the effect of ethnicity on COVID-19 clinical outcomes in both published and preprint research. We found that individuals from Asian, Black and Hispanic ethnic groups are more vulnerable to SARS-CoV-2 infection compared to those of White ethnicity. In addition, those of Black ethnicity are at higher risk of ICU admission compared to White. We found limited evidence suggesting that individuals from ethnic minority backgrounds are at increased risk of death.

Our findings suggest that the disproportionate impact of COVID-19 on ethnic minority communities is likely to be attributed to infection rather than high severity of disease. Elevated prevalence of SARS-CoV-2 in Black, Asian and Hispanic ethnic groups compared to those of White ethnicity may be attributed to a number of factors relating to increased transmission.^46,47^ This includes larger household sizes comprised of multiple generations, lower socioeconomic status and deprivation, which may increase the likelihood of living in urban or deprived areas, overcrowded households, or accommodation with shared facilities or communal areas.^48,49^ Individuals from ethnic minority backgrounds are more likely to be employed in keyworker roles, or less able to work from home, and as a result have continued to have contact with others through work or commuting.^50^ In the Bureau of Labour Statistics Current Population Survey for the year 2019, Hawkins found that Black and Asian workers were more likely to be employed in occupations with both frequent exposures to infections and proximity to others.^51^ There is also evidence that ethnic minority groups experience disproportionate rates of COVID-19 morbidity and mortality in some of these occupational groups, for example among healthcare workers.^52^ Our findings that ethnic disparities exist in mortality rates at a population level may also be attributed to the higher risk of infection, as well as inequities in access to and quality of care, including language, cost, insurance, or structural discrimination.^53^

Racism and structural discrimination may also contribute to an increased risk of transmission within ethnic minority communities.^53,54^ These processes are both complex and systemic, underpinned by unequal power relations and beliefs, and operating at individual, community, and organisational levels, resulting in stigmatisation, discrimination, and marginalisation of ethnic minorities.^55^ Within the healthcare context, this contributes to inequities in the delivery of care, barriers to accessing care, loss of trust, and psychosocial stressors.^56,57^ During COVID-19, there is evidence that ethnic minorities and migrant groups have been less likely to implement public health measures, be tested, or seek care when experiencing symptoms due to such barriers and inequities in the availability and accessibility of care,^58^ underscoring critical healthcare disparities.^3,5^

We found that minority ethnic groups continue to be under-represented in research,^59^ which is likely to be exacerbated by the same barriers that contribute to disparities in access to care and health outcomes.^53^ Amongst our pooled cohort, only 16% of patients were from non-White ethnic backgrounds. In particular, pooled adjusted risk with regards to ICU admission and death were based on a small number of studies, limiting generalisability in these outcomes from our data.

### Strengths and limitations

This is the first and only comprehensive synthesis of the existing evidence base, in which we provide a robust examination of both published peer-reviewed research and preprints reporting on COVID-19 outcomes in ethnic minority groups. Whilst the findings integrate the available data in this field, providing insight into disparities in COVID-19 outcomes in ethnic minorities in order to inform both policy and practice, our study had several limitations.

Variations across papers in relation to populations, setting, treatment context, and reporting of ethnicity and outcomes, resulted in high heterogeneity. However, this does not preclude pooling of data and is consistent with other meta-analyses on infection in diverse populations.^60,61^ Instead, we explored heterogeneity through sensitivity analyses. The analyses provide an important visualisation of the data available, and highlight the heterogeneity across the research and need to improve data collection and analysis, including greater standardisation in adjusted analyses.

Several studies may have overlapping populations.^62^ For example, we found six studies from Mount Sinai investigating mortality from COVID-19; quite possibly from the same population.^39,63–66^ We have minimised this error by excluding studies which were clearly done on the same database, though we urge greater transparency in reporting for future research.

Finally, all studies included were from the UK and USA. Whilst both these countries have ethnically diverse populations, generalisability of our findings to other countries should be cautioned, and further research should be undertaken in other country contexts and diverse income-level settings.

### Implications for future research

This research highlights the critical inconsistencies in the evidence base in categorisations of ethnicity and race, disaggregation of data, inclusion of confounders, and reporting standards. For example, only 6% (41/741) of potentially eligible articles we screened investigated ethnicity with relation to COVID-19 clinical outcomes. At least 8% (62/741) of studies may have generated outcome data, but did not provide this in their manuscript. Furthermore, of all 35 studies, only one separated Asian into South Asian, Pakistani and Chinese; an evidently heterogeneous group.^27,67^ This inhibits the ability to draw from the existing research to inform evidence-based policy and practice, and highlights the need to standardise data collection and reporting of ethnicity in research.

This research also illustrates the rapid generation of data during the pandemic, and the imperative to provide a comprehensive and transparent representation of the available evidence. As such, we included preprint articles, and did not exclude on the basis of quality. Though published peer-reviewed articles typically had higher quality scores, preprints meeting the inclusion criteria were generally of high quality (Supplementary figure 6). Ultimately, half of the studies included were preprints, underscoring the impetus to promote open access and transparency of data.

The research also highlighted the challenges in identifying emerging unpublished data in this quickly evolving evidence base. For example, two studies by Barron and colleagues and the ISARIC group^68,69^ were not included in MedRxiv. Study participants from the included OpenSAFELY publication would have significantly overlapped with Barron’s whole-population study and findings from the ISARIC publication align with our conclusions.^28^ However, this suggests the need to reimagine how preprint evidence can be disseminated and systematically examined in the post-pandemic context to enable more comprehensive and rapid synthesis of emerging findings.

### Conclusions

We found strong evidence that patients of Black, Asian and Hispanic ethnicity are more vulnerable to SARS-CoV-2, compared to those of White ethnicity. As the COVID-19 pandemic continues to evolve, more studies should aim to present outcomes(hospitalisation, ICU admission and death) of patients with COVID-19 disaggregated by ethnicity, with data adjusted by key confounders. Policy-makers should strongly consider strategies to minimise exposure risk of SARS-CoV-2 in ethnic minority groups, by facilitating access to timely care, and targeting social determinants, structural racism, and occupational risk underlying inequities.

## Data Availability

This is a meta-analysis of all published/data presented on preprint servers. Therefore the data is publicly available

## Supplementary materials

Supplementary material 1: Search strategy for meta-analysis

Supplementary material 2a: Forrest plot of pooled prevalence of SARS-CoV-2 by ethnicity

Supplementary material 2b: Forrest plot of pooled unadjusted risk of SARS-CoV-2 infection by ethnicity (Reference group: White)

Supplementary material 2c: Forrest plot of pooled adjusted risk of SARS-CoV-2 infection by ethnicity (Reference group: White)

Supplementary material 3a: Forrest plot of pooled prevalence of ICU admission for COVID-19 by ethnicity (Reference group: White)

Supplementary material 3b: Forrest plot of pooled unadjusted risk of ICU admission for COVID-19 by ethnicity (Reference group: White)

Supplementary material 3c: Forrest plot of pooled adjusted risk of ICU admission for COVID-19 by ethnicity (Reference group: White)

Supplementary material 4a: Forrest plot of pooled prevalence of death from COVID-19 by ethnicity (Reference group: White)

Supplementary material 4b: Forrest plot of pooled unadjusted risk of death from COVID-19 by ethnicity (Reference group: White)

Supplementary material 4c: Forrest plot of adjusted risk of death from COVID-19 in Black individuals (Reference group: White)

Supplementary material 4d: Forrest plot of adjusted risk of death from COVID-19 in Asian individuals (Reference group: White)

Supplementary material 4e: Forrest plot of adjusted risk of death from COVID-19 in Hispanic individuals (Reference group: White)

Supplementary material 4f: Forrest plot of adjusted risk of death from COVID-19 in Mixed individuals (Reference group: White)

Supplementary material 5: Funnel plots and Egger’s test for the risk of infection, admission to ICU and death

Supplementary table 6: Quality rating score for each study.

